# Greater Covid-19 Severity and Mortality in Hospitalized Patients in Second (Delta Variant) Wave Compared to the First: Single Centre Prospective Study in India

**DOI:** 10.1101/2021.09.03.21263091

**Authors:** Raghubir S Khedar, Kartik Mittal, Harshad C Ambaliya, Alok Mathur, Jugal B Gupta, Krishna K Sharma, Yogendra Singh, Gunjan Sharma, Akhil Gupta, Vaibhav Bhargava, Kishore Mangal, Anil K Sharma, Yatendra K Gupta, Pramod Sarwa, Bhawani S Mishra, Swati Sharma, Krishnakumar Sharma, Rajeev Gupta

**Affiliations:** Departments of Medicine, Eternal Hospital & Eternal Heart Care Centre and Research Institute, Mount Sinai New York Affiliate, Jaipur, India; Departments of Pulmonary Medicine, Eternal Hospital & Eternal Heart Care Centre and Research Institute, Mount Sinai New York Affiliate, Jaipur, India; Departments of Critical Care Medicine, Eternal Hospital & Eternal Heart Care Centre and Research Institute, Mount Sinai New York Affiliate, Jaipur, India; Departments of Clinical Research, Eternal Hospital & Eternal Heart Care Centre and Research Institute, Mount Sinai New York Affiliate, Jaipur, India; Departments of Microbiology, Eternal Hospital & Eternal Heart Care Centre and Research Institute, Mount Sinai New York Affiliate, Jaipur, India; Department of Pharmacology, LBS College of Pharmacy, Rajasthan University of Health Sciences, Jaipur, India; Academic Research Development Unit, Rajasthan University of Health Sciences, Jaipur, India

## Abstract

**Background & Objective:** Covid-19 pandemic has led to multiple waves secondary to mutations in SARS-CoV-2 and emergence of variants of concern (VOC). Clinical characteristics of delta (B.1.617.2) VOC are not well reported. To compare demographic, clinical and laboratory features and outcomes in the second Covid-19 wave in India (delta VOC) with the previous wave we performed a registry-based study.

**Methods:** Successive SARS-CoV-2 reverse transcriptase-polymerase chain reaction (RT-PCR) confirmed Covid-19 patients presenting to our Advanced Covid Care hospital were prospectively recruited. In the first phase (wave) from March-December 2020, 1395 of 7476 (18.7%) suspected patients tested positive and 863 (61.89%) hospitalized, while in second wave from January-July 2021 out of 1641 confirmed cases out of 8680 (19.4%) suspected 388 (23.6%) were hospitalized. Details of clinical and laboratory features at admission to hospital, management and outcomes in the two waves have been compared.

**Results:** In both cohorts, majority were men and 20% less than 40 years. Prevalence of hypertension, diabetes and cardiovascular diseases was more than 20%. Second wave patients had similar pre-hospitalization symptom duration but had significantly greater cough, fever and shortness of breath and lower SpO_2_ at presentation with greater lymphopenia, C-reactive proteins, interleukin-6, ferritin, lactic dehydrogenase and transaminases. In the second vs first wave patients, requirement of supplementary oxygen (47.9% vs 34.3%), prone positioning (89.2 vs 38.6%), high flow nasal oxygen(15.7 vs 9.1%), non-invasive ventilation (14.4 vs 9.5%), invasive ventilation (16.2 vs 9.5%), steroids (94.1 vs 85.9%), remdesivir (91.2 vs 76.0%) and anticoagulants (94.3 vs 76.0%) was greater (p<0.001). Median (IQR) length of stay [8 (6-10) vs 7 (5-10) days] as well as ICU stay [9 (5-13) vs 6 (2-10) days] was more in second wave (p<0.001). In-hospital deaths occurred in 173 patients (13.9%) and were significantly more in the second wave, 75 (19.3%), compared to the first, 98 (11.5%); unadjusted odds ratio (95% CI) 1.84 (1.32-2.55) which did not change significantly with adjustment for age and sex (2.03, 1.44-2.86), and age, sex and comorbidities (2.09, 1.47-2.95). Greater disease severity at presentation was associated with mortality in both the waves.

**Conclusions:** Covid-19 patients hospitalized during the second wave of the epidemic (delta variant) had more severe disease with greater dyspnea, hypoxia, hematological and biochemical abnormalities compared to first wave patients. They had greater length of stay in intensive care unit, oxygen requirement, non-invasive and invasive ventilatory support. The in-hospital mortality in the second wave was double of the first.

## INTRODUCTION

Delta variant (B.1617.2) of Severe Acute Respiratory Syndrome Novel Coronavirus-2 (SARS CoV-2) has emerged as the most widespread cause of Covid-19 in recent months.^1,2^ This variant, first isolated in India,^3,4^ has overtaken all the previous strains and has spread rapidly among both unvaccinated and vaccinated individuals globally.^2^ It is currently the dominant variant in Europe and North America and has also spread to Africa, South America and other Asian countries.^5^ Studies from UK, USA and Europe, with high proportion of vaccinated individuals, have reported that the strain leads to widespread breakthrough infections among the vaccinated.^6-11^ This variant has also led to widespread disease among the unvaccinated.^5,11^ In the initial phase of infection, it was reported that the delta variant is more transmissible but leads to less severe Covid-19 which was identical to the alpha variant induced disease.^12,13,14^ A rapid increase in case-numbers, which has been observed in most countries where the variant has spread, followed by a rapid decline was considered supportive of this hypothesis. It has now been established that virus load following the infection is high and rapid transmissibility of the strain is a major concern.^15^ Data from USA have reported that the number of hospitalizations following this infection have reached pre-delta periods.^15^ Comparative mortality data, between the present delta variant-wave and the previous waves of Covid-19, are not yet available and could be lower due to high vaccination rates in the US. Similar observations have been reported from Israel, UK and most Western European countries with high vaccination rates.^5,11,15^

In India, there have been two clear waves of Covid-19 related cases and deaths, the first wave began in mid-March 2020, peaked in July-October, with significant interstate variation, and declined significantly by December 2020.^3,4,16-18^ In the early phase of the epidemic in India, SARS-CoV-2 genome sequencing data were limited,^19^ since mid-2020’s there has been a substantial increase in this effort.^19^ In the first wave of the Covid-19 in India variants identified were combinations of wild-type viruses and D614G mutations.^3^ There was moderate increase in alpha variant (B.117) in North India in January 2021 but since March 2021 the dominant strain in India has been delta (B.1617.2) which has been responsible for more than 90% of infections.^4,16,18,19^ Anecdotal reports suggested that disease severity and mortality in the two waves of Covid-19 in India was similar.^20^ Modelled estimates based on serosurvey data predicted lower mortality in the second wave.^21,22^ There are only limited studies that have compared the clinical features and outcomes in Covid-19 patients in the first and second waves: a multisite registry by Indian Council of Medical Research,^23^ and a couple of hospital based registries.^24,25^ These studies reported different patient outcomes in the first and second waves of Covid-19. Therefore, to evaluate differences in clinical and laboratory features, management patterns and outcomes among hospitalized patients at a dedicated tertiary level Covid-19 management centre in India during the first (June-December 2020) and second (January-June 2021) waves of the infection we performed an observational study.

## METHODS

We initiated a registry of all patients with Covid-19 admitted to this hospital since April 2020. The registry protocol was approved by the institutional ethics committee (Government of India registration, CDSCO No. ECR/615/Inst/RJ/2014/RR-20). Informed consent from all the patients or next-of-kin was obtained for anonymized data publication as permitted by the ethics committee.

### Setting

This is a 220-bed tertiary care hospital with major focus on critical care and cardiovascular sciences. It was designated as Advanced Covid Care Hospital by Government of Rajasthan and more than 20% beds in general wards and intensive care units were initially reserved for Covid-19 patients. This proportion was later increased to 50% and 75% as the number of critically-ill patients in the state increased.^26^ The hospital subsidized treatment to all the admitted patients according to the state government regulations. We developed a protocol for admission so that only those patients fulfilling definite clinical criteria were hospitalized.^26^ The case-report form was updated and modified from that available at the Regional Covid-Care Hospital at Government Health Sciences University.^27,28^

### Patients

All patients presenting to medical and emergency departments with symptoms suggestive of upper respiratory infections were screened with reverse transcriptase-polymerase chain reaction (RT-PCR). Details of the sample collection and testing protocol have been reported.^28^ We obtained details of case history, vital systems monitoring for all patients were recorded and a flow chart of all in-hospital investigations maintained. All the hospitalized patients received clinical management according to the Indian national and international protocols.^29,30^ Essentially, hydration and oxygenation were maintained using intravenous fluids and nasal-cannula based oxygen supplementation as needed. Steroids were used only in those patients needing advanced non-invasive or invasive ventilatory support. Remdesivir was used according to the Indian national and US guidelines.^27.28^ Anti-interleukin drugs were used infrequently and nonevidence-based therapies such as oral hydroxychloroquine, ivermectin, anti-viral drugs or plasma therapy were not recommended for hospitalized patients.

### Statistical analyses

All the data were computerized and entered into MS-Excel work-sheets. We focused on clinical history at presentation, hematological and biochemical investigations at admission, imaging, medical and other therapies and in-hospital outcomes. Follow-up data have not been recorded. Descriptive analyses have been performed using SPSS package (Version 21.0). The categorical variables have been reported as numbers and percent while continuous variables are reported as medians and 25-75^th^ percentile interquartile range (IQR). Inter-groups comparisons have been performed using χ^2^-test for categorical variables and non-parametric Kruskal-Wallis test for continuous variables. To determine odds ratios (OR) and 95% confidence intervals (95% CI) for deaths in second vs first wave patients, we initially performed univariate logistic regression. Multivariate logistic regression was performed using variables likely to confound the outcomes such as age, sex and comorbidities. We did not adjust for disease severity and other clinical and outcome measures, as these were the primary study outcomes and likely to attenuate the OR’s and make comparisons of disease severity difficult. P values less than 0.05 are considered significant.

## RESULTS

In the first wave from June-December 2020, 7476 suspected patients were evaluated with nasopharyngeal samples for SARS-CoV-2 antigen RT-PCR test, 1395 (18.7%) tested positive for the virus and 863 individuals (61.9%) were hospitalized. In the second wave from January-July 2021, 1641 of 8680 (19.4%) suspected patients tested positive for the virus and 388 (23.6%) were hospitalized. Higher hospitalization rates in the first wave were due to government guidelines that encouraged hospital admission and isolation for all individuals with virus-positive reports,^27,29^ while lower hospitalization rates in the second wave were due to change in government guidelines that encouraged home-based isolation and also due to non-availability of beds at our hospital to some of the sicker patients.^31^

Demographic details (Table 1) show that more than two-third patients were men, although number of women was significantly greater in the second wave. One in five were younger than 40 years in both cohorts and age distribution was similar. Prevalence of comorbidities, especially hypertension, diabetes and cardiovascular diseases was high in both the cohorts. Median duration of symptomatic illness was similar in second and first waves. Second wave patients had significantly greater cough, fever and shortness of breath at presentation. This group also had significantly higher pulse rate and lower SpO_2_ at presentation (Table 1). All the Covid-19 related adverse hematological and biochemical parameters-lymphopenia, C-reactive proteins (hsCRP), interleukin-6, ferritin, lactic dehydrogenase and transaminases were more in second wave patients (Table 1).

**Table 1:** Demographic, clinical and laboratory variables in the first and second Covid-19 waves.

|  | **Total(N=1251)** | **Wave-1, (N=863)** | **Wave-2, (N=388)** | **P value** |
| --- | --- | --- | --- | --- |
| Male | 876(70.0) | 626(72.5) | 250(64.4) | 0.219 |
| Female | 375(30.0) | 237(27.5) | 138(35.6) | 0.035 |
| Age≤45 Yr | 259(20.7) | 170(19.7) | 89(22.9) | 0.291 |
| Age, 46-60 Yr | 432(34.5) | 294(34.1) | 138(35.6) | 0.719 |
| Age>60 Yr | 560(44.8) | 399(46.2) | 161(41.5) | 0.332 |
| Urban | 1123(89.8) | 779(90.3) | 344(88.7) | 0.386 |
| Rural | 128(10.2) | 84(9.7) | 44(11.3) |  |
| **Medical comorbidities** |  |  |  |  |
| Hypertension | 551(44.1) | 385(44.7) | 166(42.8) | 0.525 |
| Cardiovascular disease | 241(18.9) | 174(19.6) | 67(17.3) | 0.001 |
| Diabetes | 495(39.6) | 346(40.2) | 149(38.4) | 0.551 |
| Asthma | 39(3.1) | 23(2.7) | 16(4.1) | 0.172 |
| Chronic obstructive pulmonary disease | 33(2.6) | 26(3.0) | 07(1.8) | 0.214 |
| Other complications | 347(27.8) | 235(27.3) | 112(28.9) | 0.506 |
| **Symptoms and Signs** |  |  |  |  |
| Symptom duration (days) | 5.0 (4-7) | 5.0 (4-7) | 5.0 (3-7) | 0.542 |
| Cough | 794(63.6) | 518(60.2) | 276(71.1) | <0.001 |
| Fever | 991(79.4) | 655(76.2) | 336(86.6) | <0.001 |
| Shortness of breath | 563(45.1) | 365(42.4) | 198(51.0) | 0.005 |
| Pulse (per minute) | 88 (78-100) | 86 (78-100) | 90 (82-102) | <0.001 |
| Temperature (°F) | 98.5 (98.2-99.0) | 98.5 (98.2-98.9) | 98.6 (98.2-99.2) | 0.003 |
| Oxygen (SpO<sub>2</sub>, %) | 95 (90-97) | 96 (92-98) | 94 (89-96) | <0.001 |
| Systolic BP (mmHg) | 130 (120-144) | 130 (120-143) | 131 (120-146) | 0.249 |
| **Investigations** |  |  |  |  |
| Hemoglobin (g/dl) | 12.7 (11.4-13.9) | 12.7 (11.2-13.8) | 13.0 (11.5-14.2) | 0.017 |
| Total leucocyte count (per 10<sup>4</sup>) | 6.9 (5.0-9.8) | 6.9 (4.9-9.6) | 7.2 (5.2-10.4) | 0.318 |
| Neutrophils (%) | 79.2 (68.1-88.2) | 78.0 (66.5-86.9) | 82.8 (72.0-89.8) | <0.001 |
| Lymphocytes (%) | 15.8 (8.7-25.5) | 17.3 (9.4-27.3) | 13.2 (7.7-22.8) | <0.001 |
| Platelet count (per 10<sup>6</sup>) | 2.2 (1.7-2.8) | 2.2 (1.7-2.8) | 2.1 (1.6-2.8) | 0.114 |
| hs-CRP (mg/dl) | 6.9 (2.2-18.9) | 5.8 (1.7-15.1) | 12.2 (4.4-32.2) | <0.001 |
| d-Dimer (ng/dl) | 464 (201-982) | 511 (181-984) | 418 (226-979) | 0.005 |
| Interleukin-6 (ng/dl) | 20.1 (6.5-60.4) | 18.2 (5.1-55.6) | 24.7 (9.8-67.1) | 0.011 |
| Ferritin (mg/dl) | 351 (159-676) | 318 (148-592) | 427 (199-908) | <0.001 |
| Lactic dehydrogenase (mg/dl) | 284 (220-396) | 265 (209-375) | 308 (243-423) | <0.001 |
| Creatinine (mg/dl) | 0.94 (0.78-1.22) | 0.93 (0.77-1.20) | 0.96 (0.79-1.30) | 0.114 |
| Sodium (mEq/L) | 138 (135-141) | 139 (136-141) | 137 (134-141) | 0.001 |
| Potassium (mEq/L) | 4.5 (4.1-4.8) | 4.4 (4.1-4.8) | 4.4 (4.1-4.8) | 0.830 |
| Serum aspartate transaminase (units) | 30 (21-48) | 29 (20-44) | 35 (25-55) | <0.001 |
| Serum glutamate transaminase (units) | 28 (18-45) | 27 (18-44) | 31 (20-53) | 0.007 |
| HRCT scan thorax score (out of 25) | 12 (8-17) | 12 (8-17) | 13 (8-17) | 0.158 |
| All the continuous variables are reported as median and 25-75<sup>th</sup> interquartile range. Numbers in parentheses are percent for categorical variables and IQR for continuous variables. P values derived using $\chi^2$ test for categorical variables and Kruskal-Wallis test for continuous variables. BP blood pressure, HRCT high resolution computed tomographic scan, | | | | |

Management, clinical course and outcomes are in Table 2 and shows that in second vs first wave patients the requirement of supplementary oxygen at admission (47.9% vs 34.3%), prone positioning (both awake and sedated) (89.2 vs 38.6%), high flow nasal oxygen (15.7 vs 9.1%), non-invasive ventilation (14.4 vs 9.5%) and invasive ventilation (16.2 vs 9.5%) were significantly greater (p<0.001). Greater use of standard pharmacological therapies-steroids (94.1 vs 85.9%), remdesivir (91.2 vs 76.0%) and anticoagulants (94.3 vs 76.0%) was also observed in patients in the second wave. In the second vs first wave, median (IQR) length of stay [8 (6-10) vs 7 (5-10) days] as well as ICU stay [9 (5-13) vs 6 (2-10) days] was more (p<0.001). 2 patients developed invasive mucormycosis, both in the second wave.

**Table 2:** Clinical course, management and outcomes in the first and second waves.

|  | **Total(N=1251)** | **Wave-1, (N=863)** | **Wave-2, (N=388)** | **P value** |
| --- | --- | --- | --- | --- |
| **Oxygen status** |  |  |  |  |
| O<sub>2</sub> required | 480(38.6) | 294(34.3) | 186(47.9) | <0.001 |
| Duration (days) | 3.5±6.0 | 2.3±4.7 | 9.0±8.1 | <0.001 |
| Proning | 676(54.3) | 330(38.6) | 346(89.2) | <0.001 |
| Nasal cannula | 357(28.7) | 216(25.2) | 141(36.3) | <0.001 |
| Non-rebreather masks | 139(11.2) | 78(9.1) | 61(15.7) | 0.001 |
| Non-invasive support | 137(11.0) | 81(9.5) | 56(14.4) | 0.009 |
| Invasive ventilation | 144(11.6) | 81(9.5) | 63(16.2) | 0.001 |
| **Medicines** |  |  |  |  |
| Steroids | 1107(88.5) | 742(85.9) | 365(94.1) | <0.001 |
| Remdesivir | 1010(80.7) | 656(76.0) | 354(91.2) | <0.001 |
| Anticoagulants | 1104(88.2) | 738(85.5) | 366(94.3) | <0.001 |
| Tocilizumab/ Bevacizumab | 73(5.8) | 58(6.7) | 15(3.8) | 0.043 |
| **Hospital duration** |  |  |  |  |
| Length of stay (day) | 7.0(6.0-10.0) | 7.0(5.0-10.0) | 8.0(6.0-10.0) | 0.001 |
| Wards (days) | 6.0(3.0-8.0) | 6.0(2.0-8.0) | 7.0(6.0-8.0) | <0.001 |
| ICU (days) | 7.0(3.0-11.0) | 6.0(2.0-10.0) | 9.0(5.0-13.0) | <0.001 |
| Mucormycosis | 2(0.16) | -- | 2(0.52) | -- |
| **Outcomes** |  |  |  |  |
| Discharged to home | 1055(84.9) | 745(87.1) | 310(79.9) | 0.001 |
| Transfer to hospice care | 15(1.2) | 12(1.4) | 3(0.8) | 0.358 |
| Deaths | 173(13.9) | 98(11.5) | 75(19.3) | <0.001 |
| All the continuous variables are reported as median and 25-75<sup>th</sup> interquartile range. Numbers in parentheses are percent for categorical variables and IQR for continuous variables. P values derived using $\chi^2$ test for categorical variables and Kruskal-Wallis test for continuous variables. | | | | |

In-hospital deaths occurred in 173 patients (13.9%) and were significantly more in the second wave, 75 (19.3%), compared to the first, 98 (11.5%), (Figure 1). Important causes of death were progressive respiratory failure culminating in multi-organ dysfunction and multi-organ failure. Incidence of pulmonary embolism and acute coronary syndromes was rare (n <10). A small number of patients (15, 1.2%) requested to be transferred to home-based hospice care and 85% of patients were discharged to home-based rehabilitation, lower in the second wave (n=310, 79.9%) than in the first (n=745, 87.1%) (p<0.001). Unadjusted and multivariate adjusted OR (95% CI) for deaths in second vs first waves are shown in Figure 2. Unadjusted OR were 1.84 (CI 1.32-2.55). OR’s did not attenuate with adjustment for age (1.94, CI 1.38-2.73), age and sex (2.03, CI 1.44-2.86), and age, sex and comorbidities (2.09, CI 1.47-2.95). The ORs attenuate completely following adjustment for oxygenation and respiratory support (1.67, CI 0.83-3.13).

**Figure 1:**
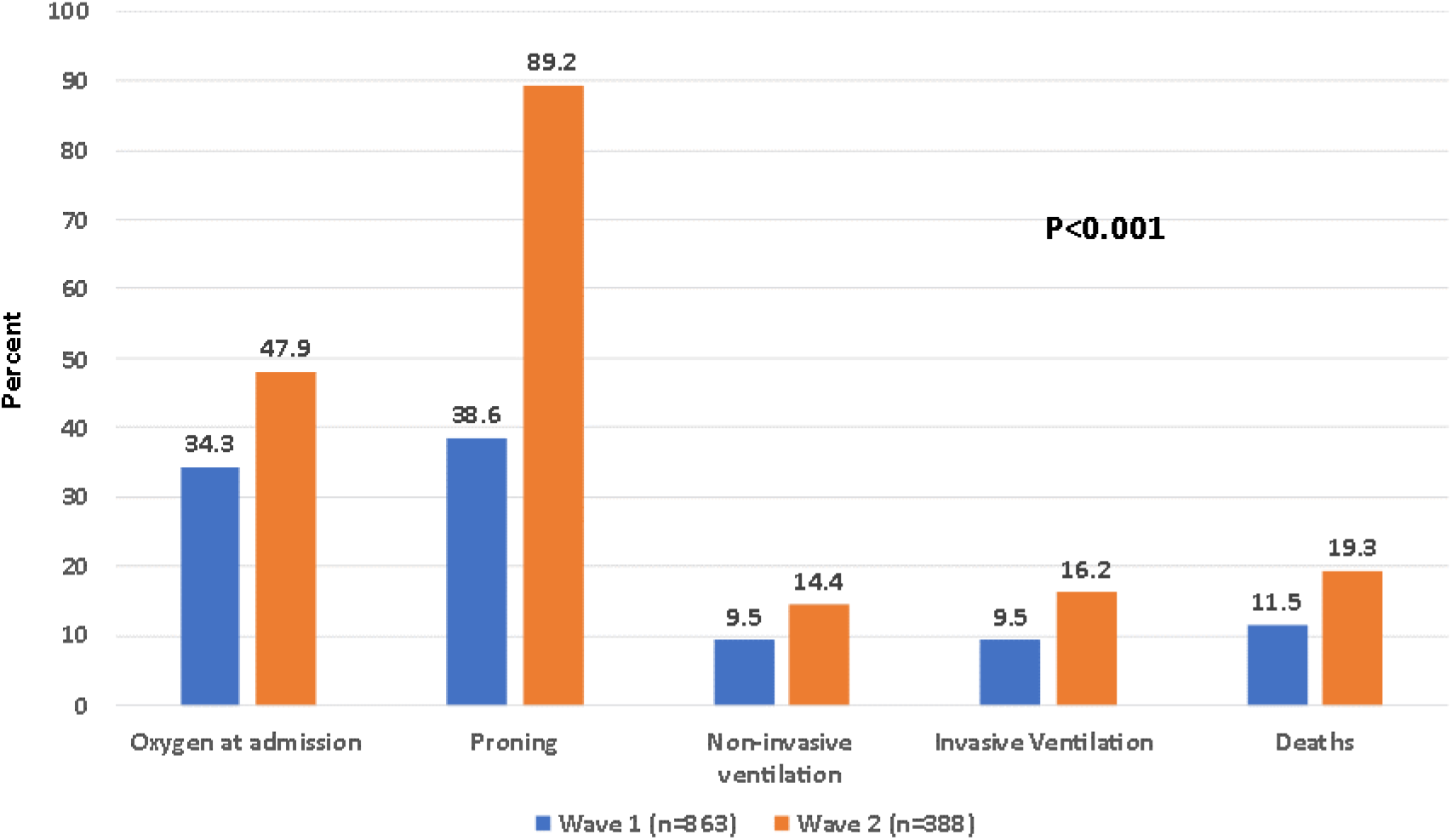
Differences in major outcomes in the two waves of Covid-19.

**Figure 2:**
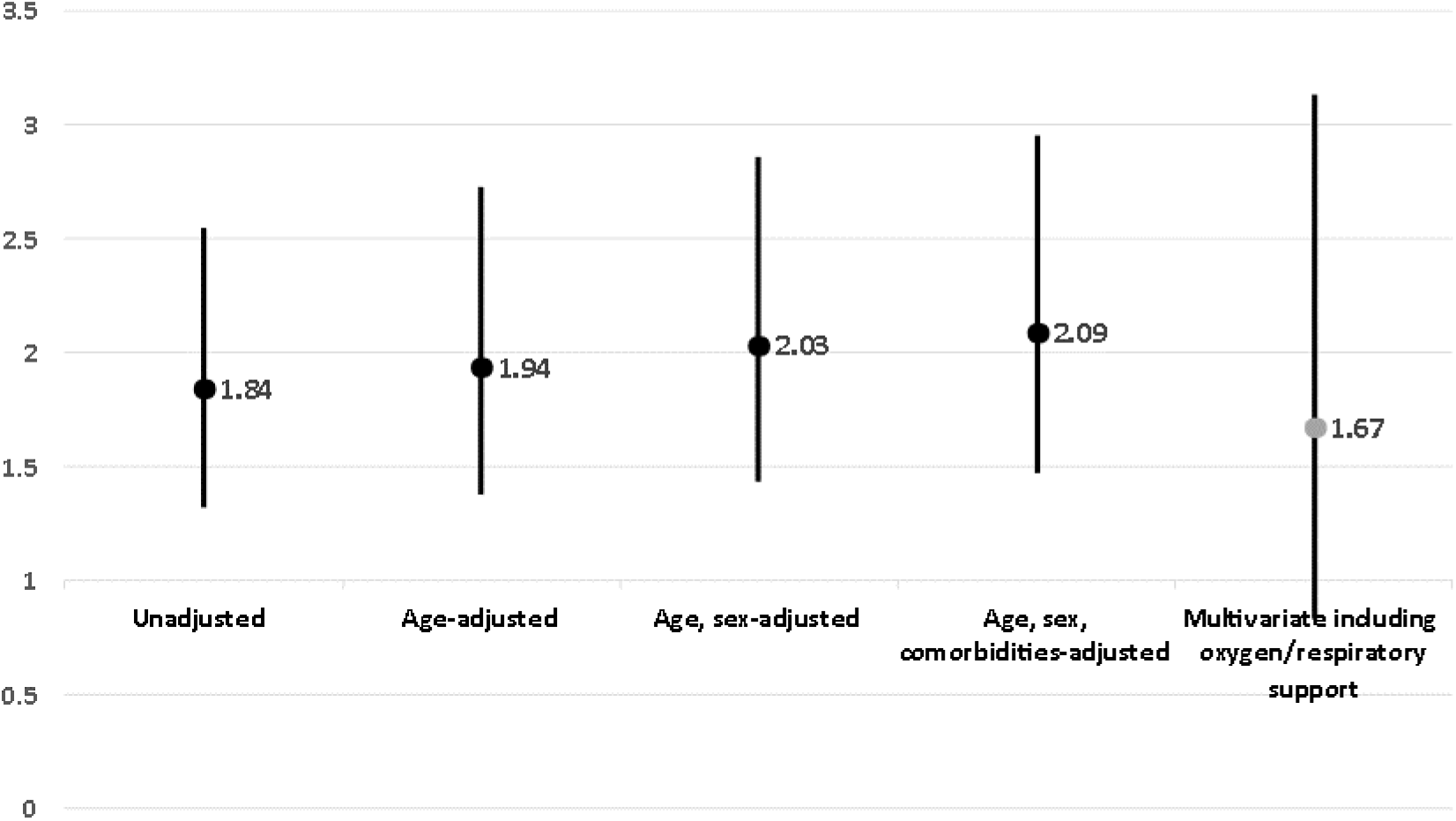
Unadjusted and Adjusted Odds Ratios (95% CI) for Deaths in Second vs First Waves.

Clinical characteristics in patients who survived and died in the first and the second waves are in Table 3. In both the waves mortality was more in men, older patients and those with hypertension, cardiovascular disease and diabetes. Patients who died were more symptomatic, had lower oxygen concentration at admission, and had more leukocytosis, lymphopenia, higher biomarkers (CRP, d-Dimer, interleukin-6, ferritin, lactic dehydrogenase and evidence of kidney and liver injury (Table 3). Length of hospital stay, oxygen requirement and non-invasive and invasive respiratory support were also more in patients who did not survive (p<0.001).

**Table 3:** Differences in Clinical Features in Covid-19 Patients Discharged or Died in the Two Waves.

|  | First wave |  | Second wave |  |
| --- | --- | --- | --- | --- |
|  | Discharged(n=745) | Deaths (n=98) | Discharged(n=310) | Deaths (n=75) |
| Male | 529(71.0) | 81(82.7)* | 198(63.9) | 49(65.3) |
| Female | 216(29.0) | 17(17.3) | 112(36.1) | 26(34.7) |
| **Co-morbid factors** |  |  |  |  |
| Age ≤45 Yr | 165(22.1) | 1(1.0)*** | 81(26.1) | 8(10.7) |
| Age 46-60 Yr | 266(35.7) | 23(23.0)** | 114(36.8) | 24(32.0) |
| Age >60 Yr | 314(42.1) | 74(75.5)*** | 115(37.1) | 43(57.3)** |
| Hypertension | 326(43.8) | 51(52.0) | 126(40.6) | 37(49.3) |
| Cardiovascular disease | 127(17.0) | 39(39.8)** | 49(15.8) | 16(21.3) |
| Diabetes | 296(39.7) | 43(43.9)* | 111(35.8) | 36(48.0)** |
| Asthma | 22(3.0) | 01(1.0) | 15(4.8) | 01(1.3) |
| COPD | 21(2.8) | 04(4.1) | 06(1.9) | 01(1.3) |
| **Clinical and biochemical features** |  |  |  |  |
| Shortness of breath, % | 284(38.1) | 72(73.5)*** | 138(44.5) | 58(77.3)** |
| Oxygen saturation (SpO<sub>2</sub>, %) | 96(94-98) | 85(70-92)** | 95(92-97) | 85(70-90)** |
| Systolic BP (mmHg) | 130(122-142) | 130(113-147) | 131(120-145) | 133(117-150) |
| Hemoglobin (g/dl) | 12.7(11.4-13.9) | 11.8(10.1-13.3)*** | 13.1(11.6-14.3) | 12.7(11.2-14.0) |
| Total leucocyte count (per 10<sup>4</sup>) | 6.5(4.8-8.9) | 11.6(7.8-14.7)*** | 6.7(5.0-9.3) | 10.4(6.9-14.0)*** |
| Neutrophils (%) | 76.5(65.2-84.4) | 90.9(83.6-93.6)*** | 79.6(69.6-87.1) | 91.2(86.5-94.7)*** |
| Lymphocytes (%) | 18.8(11.1-28.4) | 6.9(3.9-10.5)*** | 15.4(9.3-24.7) | 7.2(3.5-11.1)*** |
| Platelet count (per 10<sup>6</sup>) | 2.2(1.7-2.8) | 2.0(1.4-2.7)* | 2.1(1.6-2.8) | 1.96(1.47-2.60) |
| hs-CRP (mg/dl) | 4.6(1.4-11.3) | 27.4(15.5-67.9)*** | 9.5(3.8-26.6) | 25.9(11.6-92.3)*** |
| d-Dimer (ng/dl) | 459(167-841) | 1508(811-3329)*** | 335(192-636) | 1149(480-2353)*** |
| Interleukin-6 (ng/dl) | 15(4-42) | 124(46-338)*** | 22(8-48) | 72(16-305)*** |
| Ferritin (mg/dl) | 279(137-533) | 743(388-1629) | 369(165-694) | 1015(454-1710)*** |
| Lactic dehydrogenase (mg/dl) | 252(200-327) | 516(347-714)*** | 295(233-393) | 489(307-651)*** |
| Creatinine (mg/dl) | 0.90(0.76-1.10) | 1.46(1.06-3.48)*** | 0.93(0.77-1.18) | 1.3(0.87-1.83)*** |
| Sodium (mEq/L) | 139(136-141) | 138(135-142) | 137(134-140) | 139(134.7-142.2)* |
| Potassium (mEq/L) | 4.4(4.1-4.8) | 4.6(4.2-5.2)** | 4.4(4.1-4.7) | 4.6(4.0-5.1) |
| Serum glutamate transaminase | 27.8(19.6-40.9) | 45.7(26.7-71.3)*** | 34(23.3-52.4) | 43.0(28.9-75.8)** |
| Serum aspartate transaminase | 26.7(17.5-42.9) | 31.0(19.0-48.6) | 31.3(19.6-51.1) | 32.2(21.8-57.4) |
| HRCT scan thorax score (of 25) | 12(7-16) | 19(15-22)*** | 12(7-15) | 19(15-21.5)*** |
| **Outcome variables** |  |  |  |  |
| Length of hospital stay (days) | 7.0(6.0-9.0) | 8.0(2.7-15.0)* | 8.0(6.0-10.0) | 9.0(4.0-15.0)* |
| Oxygen required (yes/no) | 191(25.6) | 95(96.9)*** | 111(35.8) | 73(97.3)** |
| Oxygen duration (days) | 0.0(0.0-1.0) | 7.0(2.0-13.0)** | 6.0(4.0-10.0) | 8.5(4.0-15.0)* |
| Prone | 260(34.9) | 69(70.4)*** | 277(89.4) | 67(89.3) |
| Nasal cannula | 180(24.2) | 31(31.6) | 109(35.2) | 31(41.3) |
| Non-rebreather masks | 45(6.0) | 32(32.7)*** | 28(9.0) | 33(44.0)** |
| Non-invasive support | 36(4.8) | 43(43.9)*** | 20(6.5) | 36(48.0)** |
| Invasive ventilation | 7(0.9) | 74(75.5)*** | 4(1.3) | 59(78.7)** |
| All the continuous variables are reported as median and 25-75<sup>th</sup> interquartile range. Numbers in parentheses are percent for categorical variables and IQR for continuous variables. BP blood pressure, COPD chronic obstructive pulmonary disease. |  |  |  |  |
| *p<0.05, ** p<0.01, *** p<0.001, for within-group comparison |  |  |  |  |

## DISCUSSION

This study shows that Covid-19 patients hospitalized during the second wave of the epidemic in India, with predominantly delta variant of SARS-CoV-2 (Figure 3),^19^ had lower duration of symptoms and more severe disease at presentation with more dyspnea and hypoxia and more adverse hematological and biochemical abnormalities compared to the first wave patients. These patients also had greater length of stay in intensive care unit, oxygen requirement, and need for non-invasive and invasive ventilatory support. In-hospital mortality in the second wave was 2-times of the first.

**Figure 3:**
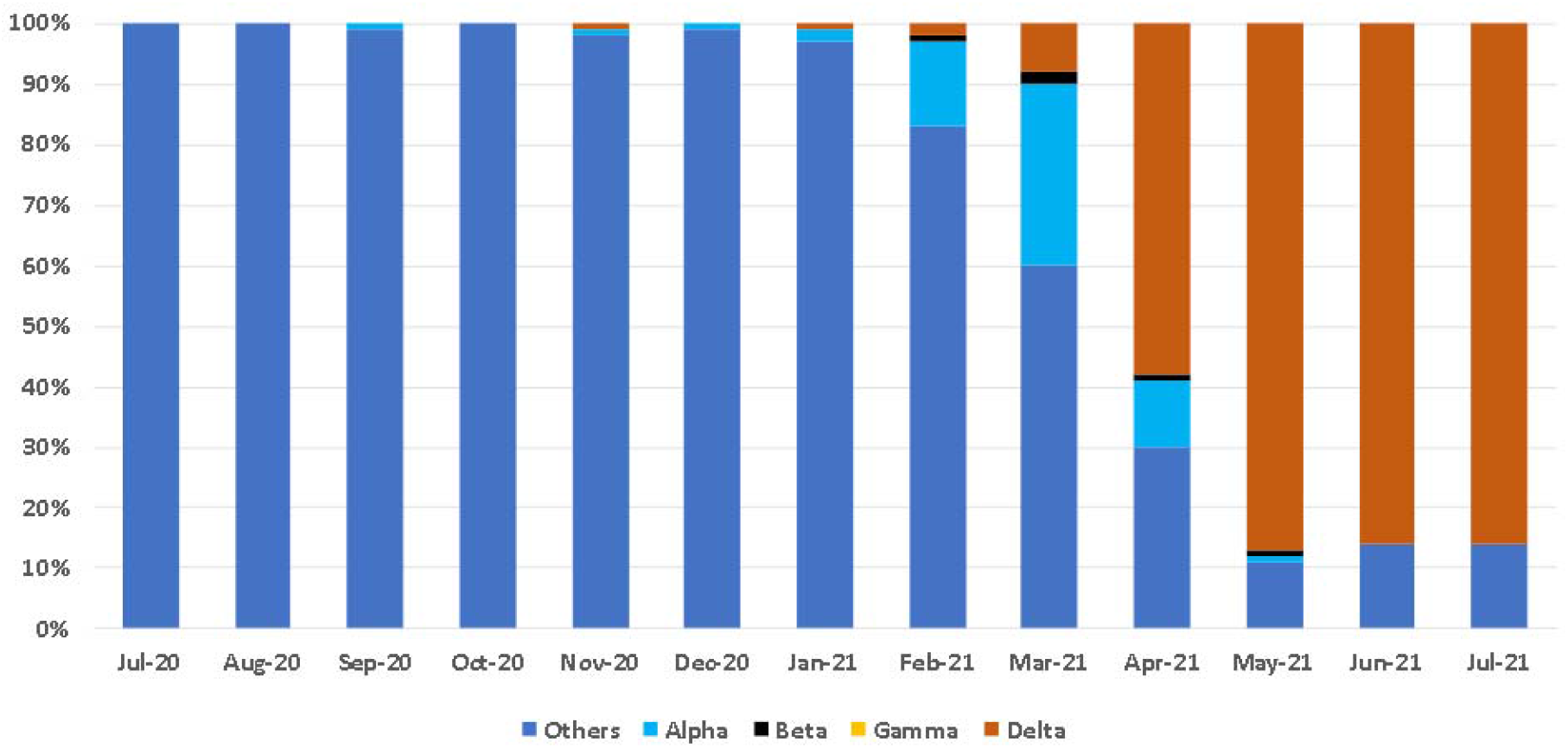
SARS-CoV-2 variants of concern in India from July 2020 to July 2021. Data source: Indian Covid-19 Genome Surveillance, INSACOG^19^.

The delta VOC of SARS-CoV-2 was first reported from India in November 2020.^3^ It is highly transmissible VOC with 60% more infectivity than the alpha variant and 100% more than the previous wild-type variants.^2^ More viral particles are found in airways of patients infected with this variant compared to others,^2^ and delta is regarded as the fastest and fittest variant so far.^32^ Studies have determined that R_o_ of delta variant is between 5 to 8, compared to 3 to 4 for alpha variant and 1.5 to 2.5 for D614G variants.^2^ Initial surveys in UK reported that symptoms of delta VOC tend to be not much different from other strains;^33^ and our study also shows that the median duration between symptoms onset and hospitalization in the second wave patients was same as in the first. Population based data from Singapore and Scotland have reported that there is a significantly greater transmission of delta VOC from index cases.^34,35^ The Scotland population-based study reported double rate of hospitalization from the virus.^35^ Another population-based cohort study in UK compared risk of hospital admission and emergency attendance for delta vs alpha variant.^36^ Researchers analyzed data from 43,338, Covid-19 patients in England from March-end to May 2021 and found that more Covid-19 patients with delta variant were hospitalized during this period than alpha variant patients with risk adjusted OR 2.26 (CI 1.32-3.89) and risk for presentation for emergency care was 1.45 (CI 1.08-1.95).^36^ The present study is not population-based and we cannot compare our results with these findings. Lesser disease severity and lower hospitalizations from delta VOC have been reported in UK and USA in vaccinated individuals.^2,9,10^ We did not obtain data on vaccination status in the second wave patients, however, vaccine penetration in India was low in the early months of vaccination program; ^17,37^ and it is likely that most of the patients hospitalized in our study were not vaccinated. This is a study limitation and should be studied in prospective studies.

There are only limited studies from India and other countries which compared clinical features and outcomes among the patients in the first and delta VOC waves. Modelling studies predicted lower mortality from the delta variant compared to the earlier VOC’s in India.^21,21^ Multisite Indian Council of Medical Research study,^23^ compared clinical characteristics and outcomes of patients in first (September 2020 to January 2021, n=12,059) and the second (February to May 2021, n=6903) waves in a nationwide multisite hospital-based observational study. The study reported greater oxygen requirement, non-invasive and invasive ventilatory support, and higher mortality (13.3% vs 10.2%, unadjusted OR 1.35, CI 1.19-1.52) among patients in the second compares to the first wave. Although clinical details were provided, which are similar to the present study, no biochemical data were published. Budhiraja et al,^24^ compared clinical characteristics and outcomes in hospitalized Covid-19 patients in a north Indian corporate chain of hospitals (n=10) in the first (n=14,398) and second (n=5454) waves. In the second wave, more patients had evidence of biomarker abnormalities and oxygen and ventilatory support although duration of hospitalization was shorter. Greater mortality was reported in the second vs first wave (10.5% vs 7.2%, +45.8%). A study in Gujarat reported similar mortality due to Covid-19 among renal transplant recipients in the second vs first wave.^25^ In this study of 259 renal-transplant recipients (first wave 157, second wave 102) there was no difference in presenting clinical and biochemical presentation while incidence of invasive mucormycosis and ICU admission was greater in second wave with insignificant differences in mortality in the two waves (9.6% vs 10.0%). In the present study we observed greater mortality in both second and first waves as compared to these Indian studies (19.3% vs 11.5%). Importantly, the incidence of mortality was double in the second wave compared to the first (OR 1.84, CI 1.32-2.55; adjusted OR 2.09, 1.47-2.95) (Figure 2). Delta variant has now spread to other Asian countries as well as to Europe and North America.^2,5,11^ Details of clinical manifestations and disease severity are now emerging. A population-wide study from Singapore compared clinical and virological features in alpha, beta and delta VOCs.^38^ Delta variant infection was associated with greater disease severity and deaths (OR 4.90, CI 1.43-30.78). Studies in Finland, UK and USA have reported high level of protection from delta VOC among healthcare workers.^33,39,40^ Thus, all these studies show that the delta variant is more virulent and is associated with significantly higher mortality compared to other VOCs.

In the present study, apart from those noted above, there are some more limitations. We have used hospital-based data to assess disease severity and mortality. In a vast country with variation prevalence of VOCs,^41^ and in reporting of mortality, this may not be the best strategy, as many serious patients may not have access to advanced hospital care in a cosmopolitan location leading to substantial underestimation of deaths.^42^ The best method to overcome this limitation is availability of carefully collated population-level data. In UK where population level data are available, rates of hospitalizations due to delta VOC are twice of the previous waves due to other variants (alpha, etc.).^36^ Higher rates of deaths in our cohort, as compared to previous local and national studies, could indicate a selection bias as our hospital is a high-end tertiary care centre (www.eternalheart.org) with expertise in complex disease management. Our mortality rates are, however, lower than the rates of 20-30% reported from New York (USA)^43^ and Lombardy (Italy)^44^ in first wave of Covid-19. A registry of 4645 patients from Rajasthan state reported mortality rate of 7.3% in the first wave,^45^ which is lower than the present study. Other limitations of the present study include lack of more detailed data on causes of deaths and data on medium and long-term outcomes of discharged patients. Previous studies have reported greater frequency of fungal infections (mucormycosis, aspergillosis, etc.) in Covid-19 in India due to delta VOC,^46^ mostly after discharge of patients from the hospital, and we observed only 2 cases (Table 2). We are in the process of collecting data on 6-12 months follow-up to identify the frequency and clinical details of Post-Acute Covid-19 Syndrome.^47^

The more virulent delta variant of SARS-CoV-2, as observed in the present study, presents a serious challenge to controlling the Covid-19 pandemic. Effectively responding to this formidable variant will require an evidence-based response including strict implementation of non-pharmaceutical interventions and vaccinations that, unfortunately, has not been the case for many countries.^48^ Vaccines are the only way forward that will preserve the health care infrastructure and the economy and eventually contain the pandemic.

## Data Availability

All the data have been provided in the manuscript.

